# Age at menopause and risk of ischemic stroke: a systematic review and meta-analysis

**DOI:** 10.1101/2023.10.23.23297444

**Authors:** Jie Wang, Ying Chen

## Abstract

**Background:** Despite its importance in being among the top 10 causes of old women death, there is limited data on age at menopause and ischemic stroke.

**Aims:** We performed a systematic review and meta-analysis to estimate the effect of age at onset of menopause on ischemic stroke.

**Methods:** We screened four databases (PubMed, Cochrane, Web of Science, and EMBASE databases) up to July 17, 2023. This systematic review was reported in accordance with the Preferred Reporting Items for Systematic Reviews and Meta-analyses (PRISMA) guidelines and was registered with PROSPERO (CRD42023444245). Data extraction and quality assessment were independently undertaken by two reviewers. A random-effects model was used for meta-analysis using Revman5.4 to calculate the risk ratio of the incidence of ischemic stroke. Heterogeneity was assessed using I^2^. Meta-regression and assessment for bias were performed.

**Results:** Out of 725 records identified, 10 studies were included in the qualitative synthesis and the quantitative meta-analysis. The pooled incidence rate for ischemic strokes which age at onset of menopause before 43 years old was 1.22 (95% confidence interval (CI): 1.02-1.46). The pooled incidence rate of early menopause was 1.26 (95% CI: 1.07-1.48) for ischemic stroke. The incidence rate of ischemic stroke for women with early menopause may be in an environment with a high incidence for a long time.

**Conclusion:** Evidence from this meta-analysis suggests that early menopause is associated with an increased risk of ischemic stroke. Age at onset of menopause before 43 years old may be the cut-off value of increased risk of ischemic stroke.

## Introduction

The risk of cardiovascular and cerebrovascular diseases (CCVD) increases with aging, and because women tend to live longer than men, more women suffer from and die from CCVD ^[1]^. Therefore, early identification of women at high risk of CCVD and timely implementation of appropriate lifestyle or therapeutic interventions are of public health importance. As the most common manifestation of cerebrovascular disease, stroke is the fourth leading cause of death and a major cause of severe disability^[2, 3]^. Ischemic stroke could result from multiple events such as embolism with a cardiac origin, occlusion of small vessels in the brain, and atherosclerosis affecting the cerebral circulation. Ischemic stroke is the most important causes of neurological morbidity and mortality^[4]^.

The risk for ischemic stroke was significantly lower in midlife women than men, but this difference diminished with age^[5, 6]^. Currently, known risk factors for stroke such as hypertension, smoking, and ischemic heart disease, are more common in men ^[7]^. However, they can only partly explain the differences in stroke incidence. Furthermore, little attention has been paid to the influence of sex-specific risk factors on the risk of ischemic stroke.

Female estrogen participates in the relaxation and dilation of blood vessels, accommodating blood flow and activating the renal angiotensin-aldosterone system. Therefore, reduced estrogen levels after menopause not only harden the blood vessels^[8]^, but also lead to endothelial dysfunction, inflammation, and immune dysfunction^[9]^. After menopause, the endogenous estrogen level decreases, and the risk of CCVD increases dramatically, making menopause as a high-risk period for CCVD in women^[10, 11]^. Within the first 10 years after menopause, a woman’s risk for ischemic stroke roughly doubles^[12]^. Therefore, the importance of cardiovascular and cerebrovascular risk assessment and prevention should be emphasized during this period.

The age at onset of menopause may be a marker not only of reproductive aging but also of general health and physical aging^[13]^. Early menopause may affect the risk of ischemic stroke, and women with early menopause may be in an environment with a high incidence of CCVD for a long time^[14]^.However, the exact relationship between age at menopause and ischemic stroke is still unclear. This article intends to focus on the effect of age at onset of menopause on ischemic stroke.

We performed a systematic review and meta-analysis of the available observational evidence to quantify the association of age at menopause with the occurrence of ischemic stroke.

## 2. Materials and methods

### 2.1. Search strategy and selection criteria

We conducted a systematic review of published articles up to July 17, 2023, using the PubMed, Cochrane, Web of Science, and EMBASE databases. The search was restricted to studies published in English and human studies using the following search keywords and medical subject heading terms in the PubMed (MeSH), Cochrane, Web of Science, and EMBASE (Emtree) : (menopause OR menopausal OR post-menopause OR postmenopause OR post-menopausal OR postmenopausal) AND (“Ischemic stroke “OR” Ischemic strokes “OR” Ischemic cerebrovascular disorder”). We further reviewed the references of all included articles to identify any additional studies not identified by our search strategy (known as the snowballing approach). This systematic review was reported in accordance with the Preferred Reporting Items for Systematic Reviews and Meta-analyses (PRISMA) guidelines and was registered with PROSPERO (CRD42023444245).

Wang and Chen screened the titles and abstracts of identified articles and reviewed the full text of relevant articles. Articles that meet Our inclusion criteria are observational studies (i.e., cohort or case-control study design) reporting age at natural menopause (surgical and hypothalamic pituitary amenorrhea are excluded) and examining the association with ischemic stroke. Studies were excluded if they were review papers, commentaries, conference abstracts, and studies without relevant exposures, outcomes, or effect estimates (i.e., relative risk, hazard ratio, or odds ratio). We contacted original authors by E-mail to obtain missing data. Excel2021 was used to tabulate and Revman5.4 was used to visually display results of individual studies and syntheses. The review protocol can be accessed from supplement files.

### 2.2. Data extraction

Data extraction and quality assessments were conducted by two independent reviewers (Wang and Chen), and any disagreement was resolved by discussion. The following information was extracted from the eligible articles: first author, date of publication(year), study title, year of baseline survey, location of study, mean age of baseline, length of follow-up(year), sample size and cases, study participants, menopausal status, categorical age at menopause, ascertainment of ischemic stroke. The multivariable-adjusted relative risks (RRs) with 95% confidence intervals (CIs) was extracted for the meta-analysis.

### 2.3. Quality assessment

The Newcastle-Ottawa Scale for observational studies was used to assess the risk of bias for the included articles. Selection, comparability, and outcome/exposure assessment were rated separately for cohort and case-control studies, and the rating scores ranged from 0 to 9 (highest to lowest degree of bias). Scores of 0-3, 4-6, and 7-9 were classified as low, moderate, and high quality, respectively. Begg’s funnel plots were adopted to examine the publication bias for all primary and secondary outcomes defined above.

### 2.4. Statistical analysis

Random-effects meta-analysis was conducted to synthesize results from the included studies. We specifically examined the risk of ischemic stroke associated with age at menopause, and separately examined the risk of ischemic stroke using early and late menopause as reference categories. All analyses were conducted using log-incidence rates and log-standard errors derived from the 95% CIs. In the meta-analysis, the distribution of incidence rates and 95% CIs obtained were examined using a forest plot. Pooled estimates of the incidence rate were used to determine an overall rate using a random-effects model. Assessment of heterogeneity was performed using the I^2^ statistic and the test of heterogeneity (Q-statistic); values of I^2^ > 50% and p < 0.05 were considered significant heterogeneity between studies^[15]^.

Subgroup analyses were conducted to explore potential causes of heterogeneity by publication year, study setting, study design and overall quality assessment score. A test of group differences (Cochran’s Q) was used to test for the presence of heterogeneity between groups.

## 3. Results

### 3.1. Identification of relevant articles

This search strategy identified 721 articles from PubMed, Cochrane, Web of Science, and EMBASE databases (Figure 1). Four additional eligible articles were identified using the snowball method (reference screening). After removing 99 duplicate articles, 626 articles were screened based on title and abstract. Of these, 13 articles met the criteria for full-text review. 3 articles are excluded, 1 article did not have estimates reported and another 2 articles did not have relevant outcome. A total of 10 articles were selected according to the outcome events. A comprehensive review of the literature was conducted according to the inclusion and exclusion criteria, and a total of 10 articles were included (based on 9 independent studies and a Mendelian randomization analysis).

**Fig. 1.**
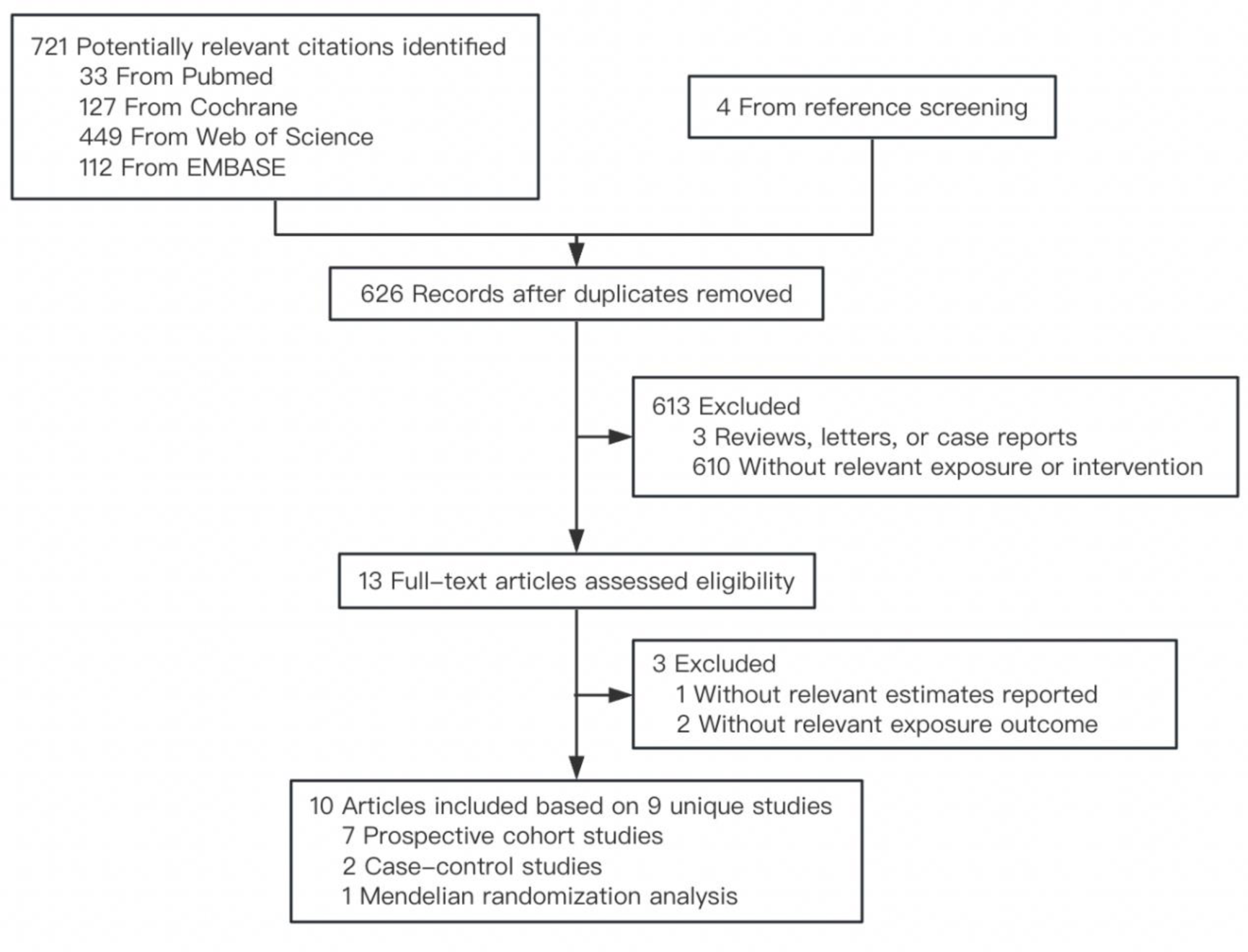
PRISMA flow diagram of the selection process and inclusion of studies.

### 3.2. Study characteristics

Of the 10 included studies, 7 were cohort studies, 2 were case-control studies, and 1 was a Mendelian randomization analysis (Table 1). Study populations were drawn from six countries (three in Asia, two in Europe, and one in the America), with the mean age of baseline ranging from 30 to 75 years (most were postmenopausal) and mean follow-up years ranging from 3 to 22 years.

**Table 1.** Characteristics of 10 included studies.

| Lead Author, Publication Date | Address of the first author | Location | Year of baseline survey | Baseline age range/average age(y) | Study design | Follow up years ( SD ) | Total participants | Outcome | Study participants | Study quality | Age at menopause (age categories) (y) | Definition of early menopause (y) |
| --- | --- | --- | --- | --- | --- | --- | --- | --- | --- | --- | --- | --- |
| Su 2023 | Department of Medicine, Seoul National University College of Medicine, Seoul, Republic of Korea. | Korea | 2009 | 66.9 ± 8.3/<br>60.8 ± 8.0 | retrospective cohort study | 8.4 | 61067/<br>1163480 | Ischemic stroke (IS) | population-based retrospective cohort study from the National Health Insurance Service database of Korea | High | < 40, 40–45, 46–50, 51–54, and ≥ 55 | <45 |
| Alonso 2007 | Department of Neurology, University Hospital Ramon y Cajal, Ctra de Colmenar Km 9,100, 28034 Madrid, Spain | Spain | 2001 | 68.9[46-93] | case-control study | 3 | 430/905 | Ischemic stroke (IS) | multicenter, age-matched case-control study conducted in 18 hospitals | High | <47, 47-50, 51-53, and ≥53 | <47 |
| Lisabeth 2009 | Department of Epidemiology (L.D.L.), University of Michigan School of Public Health, Ann Arbor, Mich | USA | 1984 | 67.2 | prospective cohort study | 22 ( 9 ) | 234/1430 | Ischemic stroke (IS) | a prospective cohort study of 5209 participants (2873 women, ages 28 to 62 at the time of enrollment) that began in 1948 in the town of Framingham, Mass | High | <42, 42-54, ≥55 | <42 |
| Hu 2021 | Department of Internal Medicine and Central Laboratory, Guangzhou | China | 2003 | 61.4 | retrospective cohort study | 12 | 222/16282 | fatal stroke | prospective cohort study including permanent Guangzhou residents aged 50 years or older that aims to examine environmental and genetic determinants of chronic diseases | High | < 43, 43–47, 48–50, 51–52, and ≥ 53 | <43 |
| Lena 2023 | Julius Center for Health Sciences and Primary Care, University Medical Center Utrecht, Utrecht, the Netherlands | Netherlands | 2006-2010/<br>1992-2000 | 58.9(SD=5.8) | Mendelian Randomization analysis | 12.6 | 6770/18909<br>5 | Ischemic stroke (IS) | UK Biobank and EPIC-CVD study | High | <40, 40-45, 45-50, 50-55, and ≥55 | <45 |
| Choi 2005 | Department of Neurology, College of Medicine, Inha University, Incheon | Korea | 1993 | 69.8±5.5 | retrospective cohort study | 5 | 93/5638 | Ischemic stroke (IS) | Korean Elderly Pharmacoepidemiologic Cohort | High | <40, 40-44, 45-49, and ≥55 | <45 |
| Michael 2019 | Cardiology Division, Massachusetts General Hospital, Harvard Medical School, Boston | USA | 2006-2010 | 40-69 | Cohort study | 10 | 9075/14426<br>0 | Ischemic stroke (IS) | adult residents of the United Kingdom recruited between 2006 and 2010. Of women who were 40 to 69 years old and postmenopausal at study enrollment | High | <40, ≥40 | <40 |
| Baba 2010 | Department of Obstetrics and Gynecology, Haga Red Cross Hospital, Tochigi, Japan | Japan | 1992-1995 | 61.0(SD=4.7) | Cohort study | 10.8( 2.2 ) | 80/3788 | Ischemic stroke (IS) | The Jichi Medical School (JMS) Cohort Study | High | <40, 40-44, 45-49, and ≥55 | <45 |
| Hu 1999 | Department of Nutrition, Harvard School of Public Health, Boston, Mass 02115, USA | USA | 1976 | 30-55 | Cohort study | 18 | 350/354326 | Ischemic stroke (IS) | The Nurses' Health Study cohort | High | <40, 40-44, 45-49, 50-54, and ≥55 | <45 |
| Hsieh 2010 | School of Public Health, Taipei Medical University | China | N/A | 66.4±9.5/<br>64.9±8.9 | case-control study | N/A | 189/192 | Ischemic stroke (IS) | People were recruited from Chi Mei Medical Center, Lotung Poh-Ai Hospital, Wan-Fang Hospital, and Taipei Medical University Hospital | High | ≤48, 49-50, ≥51 | <49 |

### 3.3. Age at menopause and risk of ischemic stroke

Table 2 shows the summary estimates from the meta-analysis of the association between age at menopause and ischemic stroke. Of these 10 studies, 5 studies defined menopausal age before 45 years old as early menopause^[16–20]^, The other 5 studies defined menopausal age before 49, 47, 43, 42, and 40 years old as early menopause, respectively^[21–25]^(Table1). For the age at menopause and the incidence of ischemic stroke, Table2B,2C,2D demonstrate the risk of menopause and ischemic stroke for women whose age at menopause was before 45, 49 and 43 years, respectively.

**Table 2.**
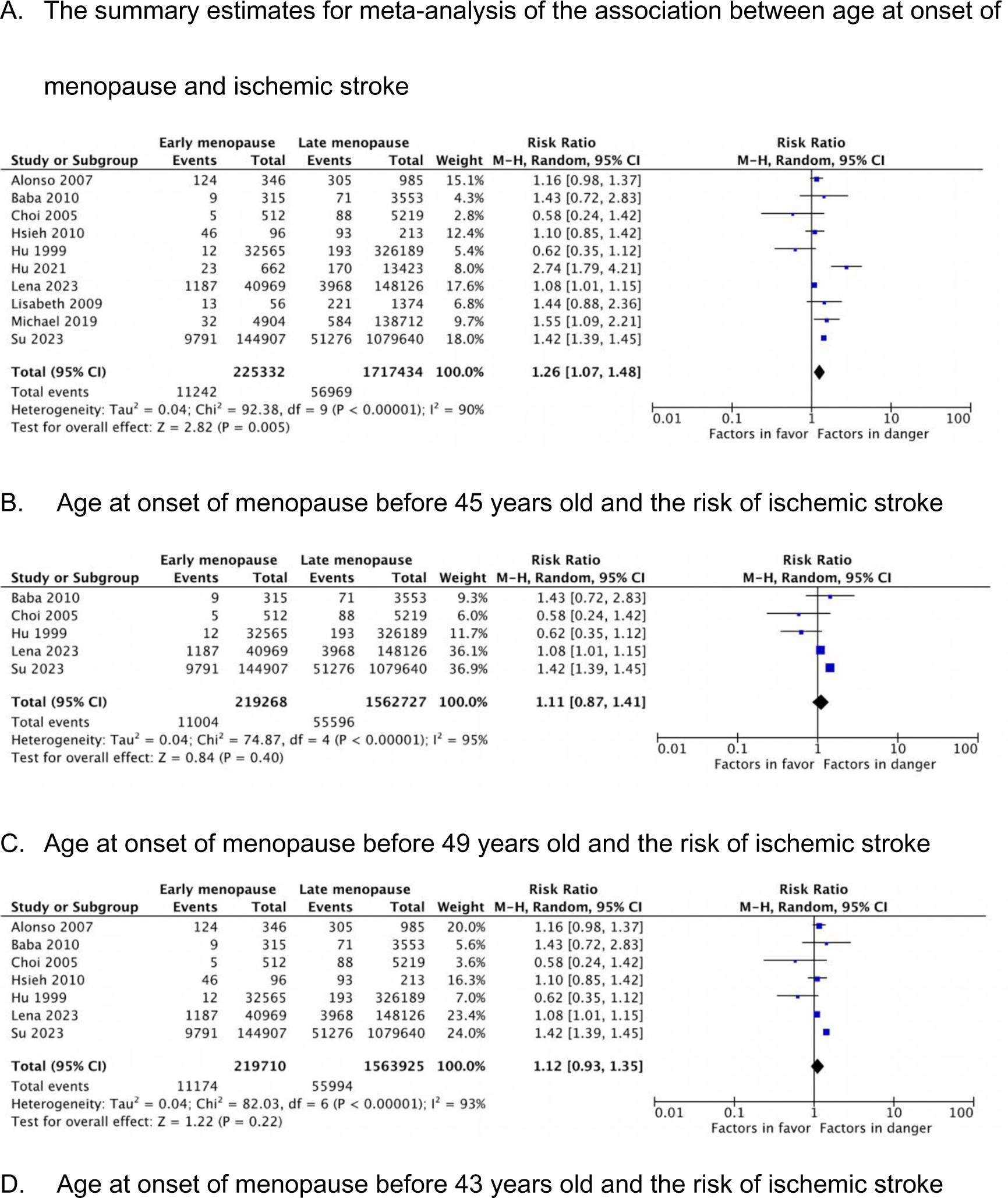
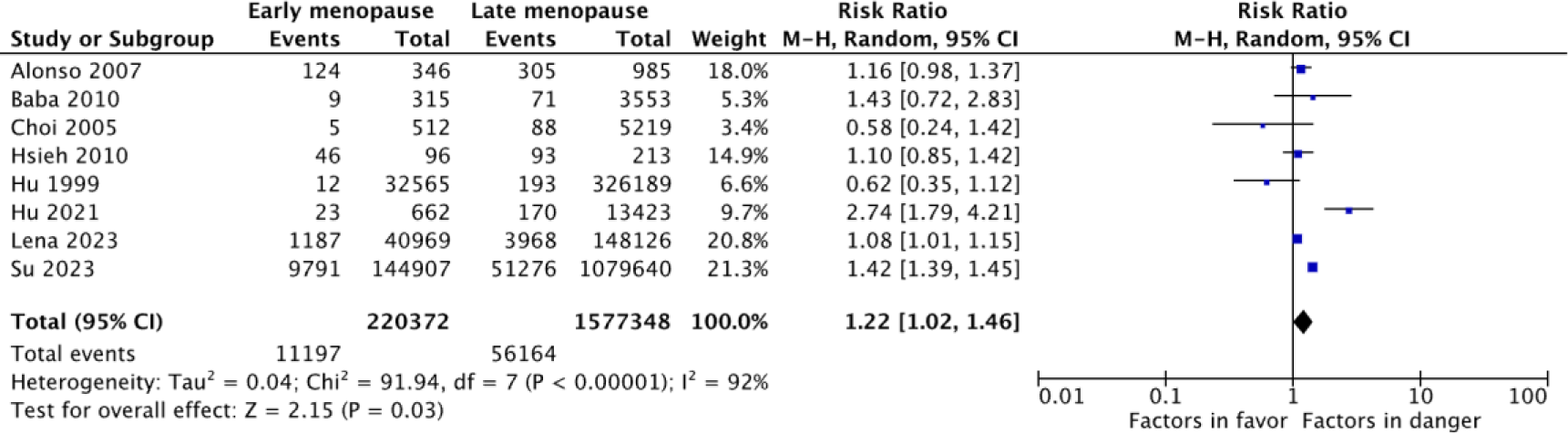
Risk of ischemic stroke for early menopausal women. A, There was evidence of between-study heterogeneity for age at menopause. Across all included studies, early menopause was associated with an increased risk of ischemic stroke (RR 1.26, 95% CI 1.07-1.48; I^2^:90%, p<0.00001). There was a significant heterogeneity between groups (p=0.005), and the group with women age at onset of menopause before 43 years contributed 24.5% to the weight. B, There was evidence of between-study heterogeneity for age at menopause younger than 45 (RR 1.11, 95% CI 0.87-1.41; I^2^:95%, p<0.00001). C, There was evidence of between-study heterogeneity for age at menopause younger than 49 (RR 1.12, 95% CI 0.93-1.35; I^2^:95%, p<0.00001). D, There was evidence of between-study heterogeneity for age at menopause younger than 43 (RR 1.22, 95% CI 1.02-1.46; I^2^:92%, p<0.00001).

There are two articles, one defines menopause age before 43 years old as early menopause^[23]^, and the other defines menopause age before 49 years old as early menopause^[26]^. Considering that there are cases of premature menopause due to iatrogenic factors or delayed menopause due to the use of hormone replacement therapy in the age at menopause group between 43 and 49 years old, there may be both early menopause and delayed menopause relative to the body’s physiological menopause age. Therefore, we considered both the population of age at menopause before 43 years old and 49 years old as early menopause groups and analyzed the heterogeneity of the articles (Table 2C, Table 2D).

Age at onset of menopause before 45 or 49 years old was not associated with an increased risk of ischemic stroke(RR 1.11, 95% CI 0.87-1.41; I^2^:95%, p<0.00001; RR 1.11, 95% CI 0.90-1.37; I^2^:94%, p<0.00001, respectively). Age at onset of menopause before 43 years old was associated with an increased risk of ischemic stroke(RR 1.22, 95% CI 1.02-1.46; I^2^:92%, p<0.00001).

### 3.4. Sensitivity analysis and publication bias

Funnel plots for the risk of ischemic stroke by menopausal age are shown in Fig. 2.

**Fig. 2.**
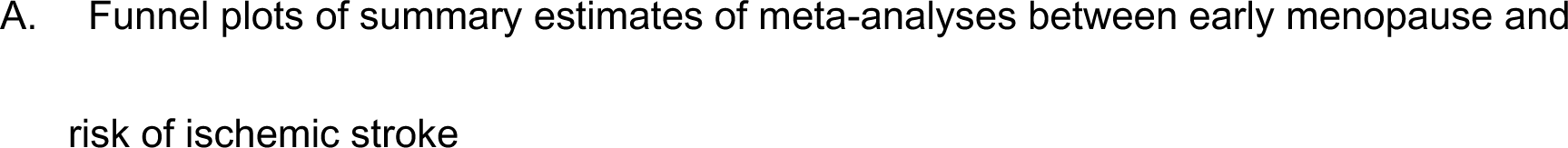
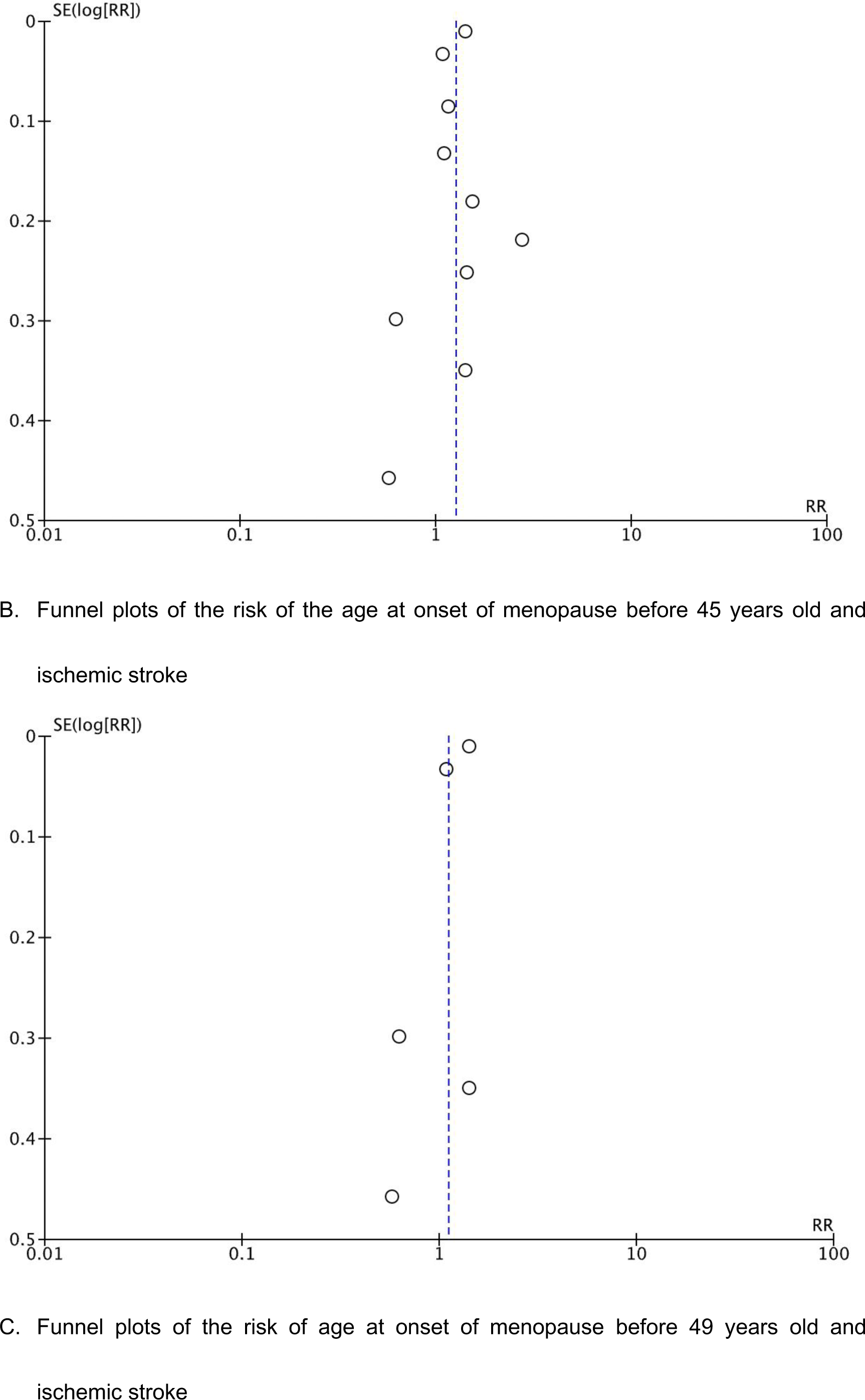
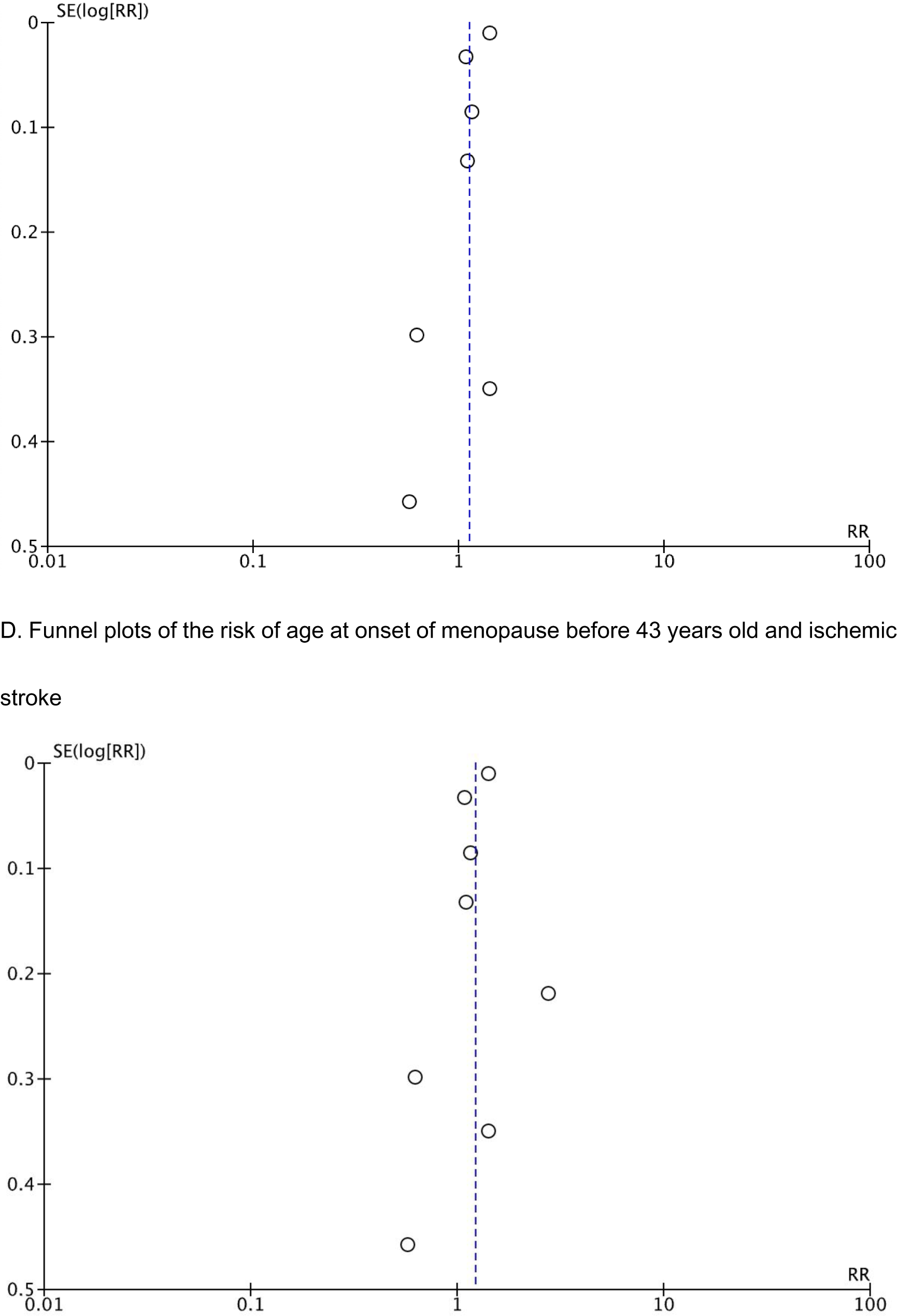
Funnel plots of risk of ischemic stroke for early menopausal women

## 4. Discussion

Relevant scholars believe that ischemic stroke is quite rare in premenopausal women, but the risk increases with age^[27]^. Menopause refers to the permanent cessation of menstruation, which represents a permanent estrogen deficiency. Studies of the association between age at onset of menopause and risk of ischemic stroke are rare and somewhat inconsistent in findings. Hu et al.^[20]^ found in the Nurses’ Health study that the age at natural menopause was not associated with the risk of ischemic stroke in individuals who had never used hormone therapy. In a cohort study of postmenopausal Korean women who had never used hormone therapy, Choi et al.^[18]^found no association between age at natural menopause and the risk of ischemic stroke, On the contrary, observational studies^[28]^ have shown that compared with men of the same age, young, premenopausal women are less susceptible to ischemic stroke, and this protection may fade away with age. Using data from the Framingham Heart Study, Lisabeth et al. ^[24]^found that women who had natural menopause before 42 years old had twice the risk of ischemic stroke compared with women who had natural menopause at the age of 42 and older. Results from a Japanese cohort study ^[19]^also suggest that after adjusting for age and risk factors, women who experienced menopause before 40 years old were more than twice as likely to have ischemic stroke as women who experienced menopause between age of 50 and 54, however, this study included both natural and surgical menopausal women.

For Chinese women, the average age to enter perimenopause is 46 years old, the average age at onset of menopause is 48-52 years old, and about 90% of women go through the menopause between the ages of 45 and 55. Early menopause is defined as menopause between the ages of 40 and 45 ^[29]^. Although the vast majority of women will follow the law of natural ovarian aging, there are still a few women who experience a decline in ovarian function decline or even failure before the age of 40. About 1% women underwent premature menopause before the age of 41, 3% women underwent menopause between the age of 41 and 46^[30]^. Early menopause can affect all systems of women, but for the occurrence of diseases in different systems, the effects of different age at menopause are also different. Muka et al.^[31]^ showed that women whose age at menopause before 45 have a higher risk of coronary heart disease, cardiovascular disease mortality, and total mortality.

For the occurrence of ischemic stroke, different from the established concept of early menopause in people whose age at menopause before 45 years old, our study showed that people whose age at menopause before 43 years old have higher risk. There was no difference in the incidence of ischemic stroke between those whose age at onset of menopause before 45 years old and those whose onset of menopause before 49 years old. This suggests that the age at onset of menopause before 43 years old is a more accurate cut-off value that reflects the relationship between menopause and ischemic stroke. There may be somewhat pathophysiological changes between 43 and 45 years of age in early menopausal women that make the different risk of ischemic stroke, which is worthwhile to further investigate the mechanism.

While this study systematically reviewed 10 primary studies concerning the incidence of ischemic stroke in early menopausal women, using recommended practice guidelines for selecting studies, it is not without limitations. The literature search was limited to studies reported in English. While this is a common practice in systematic reviews, we cannot rule out the possibility that relevant studies may have been missed due to this restriction. Owing to differential categorization of age at onset of menopause in various studies and the few studies evaluating a particular outcome (eg, ischemic stroke), some of our analysis are based on a small number of studies. Therefore, these results need to be interpreted with caution, particularly those that yielded moderate between-study heterogeneity estimates. Furthermore, the lack of studies evaluating risk in relation to time since onset of menopause limited us from performing any meaningful quantitative synthesis using this exposure.

While acknowledging these limitations, this study has some notable strengths. It is the first study to comprehensively examine the incidence of ischemic stroke in early menopausal women.

## 5. Conclusions

Evidence from this meta-analysis suggests that early menopause is associated with an increased risk of ischemic stroke. Age at onset of menopause before 43 years old may be the cut-off value of increased risk of ischemic stroke, which provides some nexus between the relationship of early menopause and ischemic stroke risk.

## Data Availability

All data in this article are available

## Availability of data, code and other materials

The data extraction for this review can be requested on a reasonable basis from a qualified researcher.

## Supplemental material

Supplemental material for this article is available online

